# Resting-State EEG Reveals Regional Brain Activity Correlates in Alzheimer’s and Frontotemporal Dementia

**DOI:** 10.1101/2024.08.05.24311520

**Authors:** Ali Azargoonjahromi, Hamide Nasiri, Fatemeh Abutalebian

## Abstract

Resting-state EEG records brain activity when awake but not engaged in tasks, analyzing frequency bands linked to cognitive states. Recent studies on Alzheimer’s disease (AD) and frontotemporal dementia (FTD) have found a link between EEG activity, MMSE scores, and age, though some findings are conflicting. This study aimed to explore EEG regional differences among AD and FTD, thereby improving diagnostic strategies. We analyzed EEG recordings from 88 participants in OpenNeuro Dataset ds004504, collected at AHEPA General Hospital using a Nihon Kohden 2100 EEG device. The study used preprocessed recordings, classification algorithms, and cognitive function assessments (MMSE) to identify significant predictors and correlations between EEG measures and cognitive variables. The study revealed that cognitive function, age, and brain activity show distinct relationships in AD and FTD. In AD, MMSE scores significantly predicted brain activity in regions like C3, C4, T4, and Fz, with better cognitive performance linked to higher EEG power in frontal and temporal areas. Conversely, age had a major influence on brain activity in FTD, particularly in regions like C3, P3, O1, and O2, while MMSE scores did not significantly predict brain activity. In FTD, higher EEG power in regions like P3, P4, Cz, and Pz correlated with lower cognitive function. Thus, the findings suggest that EEG biomarkers can enhance diagnostic strategies by highlighting different patterns of brain activity related to cognitive function and age in AD and FTD.

## 1. Introduction

Resting-state EEG (electroencephalography) records brain activity when an individual is awake but not engaged in specific tasks, analyzing delta, theta, alpha, beta, and gamma frequency bands linked to various cognitive and physiological states [1–3]. It uses electrodes placed on the scalp according to the international 10-20 system (e.g., Fp1, Fp2, F7, F3, Fz, F4, F8, T3, C3, Cz, C4, T4, T5, P3, Pz, P4, T6, O1, O2) to detect microvolt-range electrical signals from neurons. These signals are amplified and analyzed to study brain activity patterns, aiding in neurological diagnosis and monitoring changes over time. Indeed, this non-invasive approach—resting-state EEG—is used to study the brain’s baseline neural activity and can provide valuable insights into various neurological and psychiatric conditions [4–6].

In recent times, the use of resting-state EEG for studying neurodegenerative diseases like Alzheimer’s disease (AD) [7–10] and frontotemporal dementia (FTD) [11–13] has gained prominence. Indeed, it provides insights into the brain’s electrical activity patterns associated with these conditions, potentially revealing biomarkers that aid in diagnosis and monitoring disease progression. Studies have shown that EEG can detect abnormalities in frequency bands alpha power and altered connectivity patterns in patients with AD and FTD compared to healthy individuals. These findings contribute to understanding how these diseases affect brain function and may help in developing new diagnostic tools and treatment strategies [14–16].

The Mini-Mental State Examination (MMSE) is a widely used tool for assessing cognitive function and screening for cognitive impairment. It evaluates various cognitive domains, including orientation, attention, memory, language, and visuospatial skills. Research has shown significant correlations between MMSE scores and EEG activity at specific electrodes in patients with AD and FTD. These correlations help identify how cognitive functions are linked to brain activity patterns in these neurodegenerative diseases [17, 18].

In AD and FTD patients, frontal electrodes (Fp1, Fp2, F7, F3, Fz, F4, F8) have been found to correlate with MMSE scores, where lower scores are often associated with reduced EEG activity in these regions. These regions are crucial for executive functions, attention, and working memory, which are assessed by several MMSE tasks [19]. Similarly, temporal electrodes (T3, T4, T5, T6) show significant correlations with MMSE scores due to their involvement in memory and language functions [20]. Other EEG electrodes such as parietal (P3, Pz, P4), occipital (O1, O2), and central (C3, Cz, C4) electrodes are also linked to MMSE scores, with activity in these regions correlating with cognitive functions such as visuospatial skills, calculation, visual processing, and motor control. In most studies, reduced activity in these areas is associated with lower MMSE scores, indicating potential cognitive decline [21–23].

Another factor is age, which plays a crucial role in modulating brain activity and cognitive function [24]. Aging can impact EEG patterns, leading to changes in frequency band power and connectivity that are distinct from those observed in neurodegenerative diseases [25–27]. In both AD and FTD, age-related changes in brain activity can complicate the interpretation of EEG findings. For instance, age-related atrophy and functional decline can mask or exacerbate the abnormalities typically associated with these conditions [28, 29]. Noteworthy, studies have shown that age-related effects on EEG include decreased alpha and beta power and altered connectivity patterns, which can be seen across various brain regions. These changes may overlap with or confound the pathological changes observed in AD and FTD, making it important to consider age when interpreting EEG data in these populations [30–32].

However, other research finds that age-related changes in brain activity are not always consistent across different neurodegenerative conditions. For instance, in AD, age may interact with disease-specific pathology, leading to complex and sometimes conflicting results about how age influences cognitive function and brain activity compared to other conditions or in combination with other factors like genetic predispositions and lifestyle [33–36].

Despite these findings, EEG recordings can still be influenced by patient population heterogeneity, technical variability, inconsistent electrode placement, and comorbid conditions. The current study on EEG recordings addressed these limitations by including participants with AD and FTD and using standardized EEG protocols in a clinical setting. Differences in cognitive assessments and the absence of longitudinal data pose challenges to tracking disease progression accurately. To minimize technical variability, the current study implemented meticulous electrode placement, signal quality checks, and robust preprocessing techniques, including Butterworth filtering, artifact removal via ASR and ICA, and baseline correction.

The study aimed to explore the correlation between EEG measures across brain regions, cognitive performance, and age in AD and FTD patients, aiming to identify EEG biomarkers to enhance diagnostic strategies for neurodegenerative diseases.

## 2. Methods and Materials

The data presented in this article originates from the OpenNeuro Dataset ds004504, accessible via DOI: https://doi.org/10.18112/openneuro.ds004504.v1.0.7. As detailed by [37], this dataset encompasses EEG resting state recordings with eyes closed from a total of 88 participants. Within this cohort, 36 individuals were diagnosed with AD, 23 with FTD, and 29 were classified as CN. In the current study, we analyzed the data obtained from individuals with AD and FTD.

### 2.1. EEG Recording

The dataset recordings were conducted to explore functional differences in EEG activity among AD, CN, and FTD groups. These recordings were carried out in a clinical routine setting at the 2nd Department of Neurology, AHEPA General Hospital of Thessaloniki, by a team of experienced neurologists. Using a Nihon Kohden 2100 clinical EEG device, recordings were made with 19 scalp electrodes (Fp1, Fp2, F7, F3, Fz, F4, F8, T3, C3, Cz, C4, T4, T5, P3, Pz, P4, T6, O1, and O2) and 2 electrodes (A1 and A2) placed on the mastoids for impedance checking and as reference electrodes. The electrodes followed the 10–20 international system. (Figure 1)

**Figure 1.:**
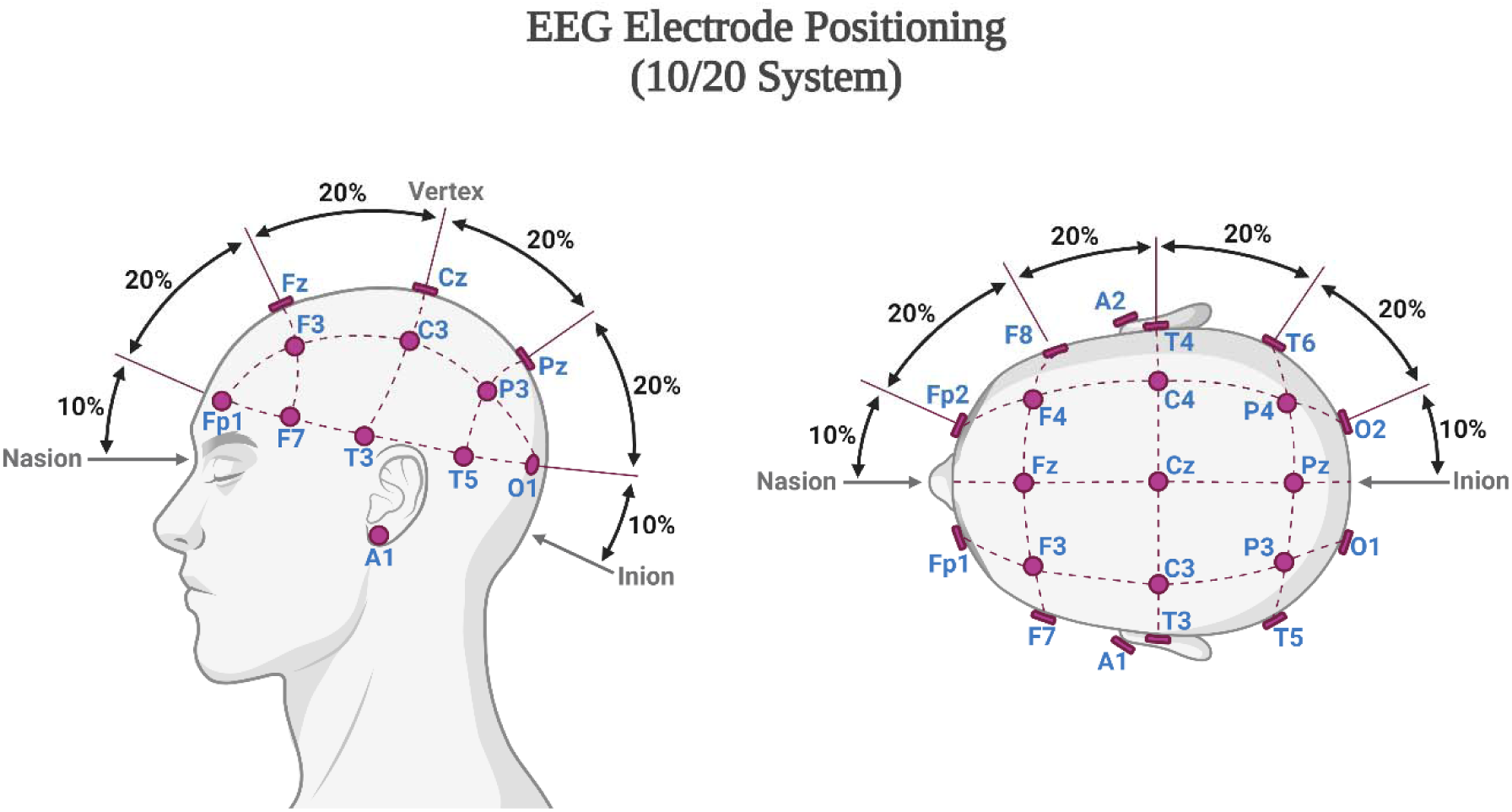
EEG electrodes positioning in followed the 10–20 international system.

Recordings adhered to the clinical protocol, with participants sitting with their eyes closed. The referential montage used Cz for common mode rejection, a sampling rate of 500 Hz, and a resolution of 10 µV/mm. The recording durations varied: 13.5 minutes on average for the AD group (range: 5.1–21.3 minutes), 12 minutes for the FTD group (range: 7.9–16.9 minutes), and 13.8 minutes for the CN group (range: 12.5– 16.5 minutes). In total, the dataset comprises 485.5 minutes of AD recordings, 276.5 minutes of FTD recordings, and 402 minutes of CN recordings.

This study received approval from the Scientific and Ethics Committee of AHEPA University Hospital, Aristotle University of Thessaloniki (protocol number 142/12-04-2023). The investigations conformed to the principles outlined in the Declaration of Helsinki (1975), revised in 2008 (http://www.wma.net/en/30publications/10policies/b3/, accessed March 2019).

### 2.2. Preprocessing

The preprocessing of the EEG signals in this study focused exclusively on the derivatives folder, where the preprocessed data is stored. Initially, a Butterworth band-pass filter with a frequency range of 0.5 to 45 Hz was applied to the signals. Following this, the signals were re-referenced to the average value of electrodes A1 and A2. To handle artifacts, the ASR (Automatic Subspace Reconstruction) routine was employed to remove segments of data that exceeded a conservative threshold, specifically a 0.5-second window with a standard deviation of 17. Subsequently, Independent Component Analysis (ICA) was used, specifically the RunICA algorithm, to decompose the 19 EEG signals into 19 ICA components [38]. Components identified as “eye artifacts” or “jaw artifacts” by the ICLabel method in the EEGLAB platform were automatically excluded. Despite the recordings being in a resting state with participants’ eyes closed, eye movement artifacts were still detected in some EEG recordings. Figure 2 in the original document visually demonstrates the difference between a raw signal and its preprocessed counterpart, highlighting the removal of high-frequency artifacts and the application of baseline correction.

### 2.3. Classification Benchmark

To benchmark the classification performance of the EEG dataset for distinguishing between AD vs. CN and FTD vs. CN, a set of straightforward and reproducible feature extraction and classification techniques were employed. This approach ensures that the methods can be easily validated and extended by other researchers.

#### 2.3.1. Feature Extraction

One of the primary features extracted for EEG classification tasks is the Relative Band Power (RBP) across five key frequency bands [39]: Delta (0.5–4 Hz), Theta (4–8 Hz), Alpha (8–13 Hz), Beta (13–25 Hz), and Gamma (25–45 Hz). The EEG signals were divided into 4-second epochs with a 50% overlap, and each epoch was labeled as AD, FTD, or CN. The Power Spectral Density (PSD) for each frequency band was calculated using the Welch method [40], which involves segmenting the signal, computing the squared magnitude of the discrete Fourier transform for each segment, and averaging the results. The relative PSD for each band was then computed, resulting in a feature matrix with five features per epoch. Figure 3 illustrates scalp heatmaps of PSD averaged across the AD, FTD, and CN groups, providing a visual comparison of the PSD distribution for each frequency band.

#### 2.3.2. Classification

To classify the EEG data for AD vs. CN and FTD vs. CN, several machine learning algorithms were utilized. These included LightGBM, Multilayer Perceptron (MLP), Random Forests, Support Vector Machine (SVM), and k-Nearest Neighbors (kNN). The classification performance was evaluated using the Leave-One-Subject-Out (LOSO) validation method [39]. In this method, all epochs from one subject are used as the test set, while epochs from the remaining subjects form the training set. This process is repeated iteratively for each subject, and the average performance metrics—accuracy, sensitivity, specificity, and F1 score—are calculated from the confusion matrix. This comprehensive benchmarking provides a solid foundation for evaluating and comparing the performance of various classification techniques on this EEG dataset.

### 2.4. Cognitive Assessment

The cognitive and neuropsychological status of the participants was assessed using the international MMSE [41]. This test evaluates various cognitive domains, including arithmetic, memory, orientation, language, and visuospatial skills. During the assessment, participants are asked a series of questions and given simple tasks to perform, such as naming objects, recalling a list of words, following basic instructions, and copying a design. The MMSE score ranges from 0 to 30, with higher scores indicating better cognitive function and lower scores indicating more severe cognitive decline.

### 2.5. Statistical Analysis

We used Python 3.11 to conduct a comprehensive analysis of EEG data from patients with AD and FTD. Our approach involved perform linear regression analyses, examining the relationships between log-transformed EEG measures and cognitive variables such as the MMSE, age, and gender. We assessed the impact of these variables on various EEG electrode sites by calculating adjusted R-squared values, beta coefficients, standard errors, 95% confidence intervals, and p-values. A p-value of less than 0.05 was considered statistically significant. Additionally, we employed correlation analysis to explore the strength of relationships between the MMSE scores and EEG measures, reporting p-values to determine statistical significance. This analysis enabled us to identify significant predictors and correlations, providing insights into how cognitive and demographic factors influence EEG patterns in AD and FTD patients.

## 3. Results

In the current study, participants were categorized into two groups: AD and FTD. The AD group consisted of individuals diagnosed with AD without additional dementia-related comorbidities. The average age of participants was 66.4 years (SD = 7.9) for the AD group and 63.6 years (SD = 8.2) for the FTD group. There was no significant age difference between the groups. The analysis revealed a notable difference in gender distribution, with a higher proportion of females in the AD group (66.7%) compared to the FTD group (39.1%). EEG measurements indicated a significant difference in log power at the O2 electrode, with FTD patients showing lower values than their AD counterparts. All participant data was anonymized in accordance with GDPR regulations to ensure the confidentiality and privacy of personal information. (Table 1)

**Table 1.:** Demographic characteristics of the participants.

|  | AD (n= 36) | FTD (n= 23) | P value |
| --- | --- | --- | --- |
| Age | 66.39± 7.89 | 63.95± 8.22 | 0.206 |
| Gender, F | 24 (66.7) | 9 (39.1) | **0.038** |
| MMSE | 17.75± 4.5 | 22.17± 2.64 | **<0.001** |
| Log Fp1 | -18.34± 2.48 | -18.86± 2.31 | 0.42 |
| Log Fp2 | -18.53± 2.48 | -18.93± 2.18 | 0.525 |
| Log F3 | -17.66±2.48 | -17.95± 2.37 | 0.655 |
| Log F4 | -17.94± 2.47 | -17.58± 2.43 | 0.586 |
| Log C3 | -18.06± 2.47 | -18.88± 2.09 | 0.19 |
| Log C4 | -17.96± 2.48 | -18.58± 2.43 | 0.346 |
| Log P3 | -18.30± 2.52 | -18.82± 2.13 | 0.411 |
| Log P4 | -18.22± 2.52 | -19.09± 2.25 | 0.184 |
| Log O1 | -18.12± 2.46 | -18.63± 2.45 | 0.434 |
| Log O2 | -17.80± 2.52 | -19.08± 2.15 | **0.049** |
| Log F7 | -18.02± 2.62 | -18.19± 2.51 | 0.816 |
| Log F8 | -18.28± 2.62 | -18.25± 2.51 | 0.965 |
| Log T3 | -18.02± 2.52 | -18.19± 2.39 | 0.792 |
| Log T4 | -18.06± 2.56 | -18.91± 2.47 | 0.215 |
| Log T5 | -18.30± 2.49 | -18.39± 2.56 | 0.894 |
| Log T6 | -17.91±2.43 | -18.45± 2.62 | 0.422 |
| Log Fz | -17.74± 2.48 | -18.13± 2.04 | 0.53 |
| Log Cz | -18.14± 2.48 | -18.22± 2.16 | 0.903 |
| Log Pz | -18.20± 2.47 | -18.44±2.18 | 0.696 |

AD: Alzheimer's Disease; FTD: Frontotemporal Dementia; MMSE: Mini-Mental State Examination.

We then employed a correlation coefficient among various EEG log-transformed power measurements and MMSE scores for individuals with AD. The correlations reflect how different EEG power measurements relate to each other and to cognitive function as assessed by the MMSE. Significant correlations are indicated by asterisks, with * representing p < 0.05 and ** representing p < 0.01.

The MMSE scores show positive correlations with several EEG regions, particularly Log Fp1, Log Fp2, Log F4, Log T3, Log T4, and Log Fz. These correlations, ranging from 0.361 to 0.379, suggest that higher cognitive function, as measured by the MMSE, is associated with increased EEG power in these areas. Notably, Log Fp1, Log T4, and Log Fz have significant correlations with MMSE scores (p < 0.05), indicating that better cognitive performance is related to higher EEG power in these regions.

In terms of EEG power measures, the correlations are predominantly strong and significant across various brain regions. For example, Log Fp1 shows robust positive correlations with Log Fp2 (0.764**), Log F3 (0.698**), and Log C3 (0.731**), highlighting a strong interrelationship among different EEG regions. These high correlations suggest a coordinated pattern of EEG activity across the brain, which is consistent in AD patients. (Table 2) (Figure 1)

**Table 2.:** A correlation coefficient between various EEG electrodes and MMSE scores for individuals with AD.

|  | 1 | 2 | 3 | 4 | 5 | 6 | 7 | 8 | 9 | 10 | 11 | 12 | 13 | 14 | 15 | 16 | 17 | 18 | 19 | 20 |
| --- | --- | --- | --- | --- | --- | --- | --- | --- | --- | --- | --- | --- | --- | --- | --- | --- | --- | --- | --- | --- |
| 1. MMSE | 1.000 | 0.214 | 0.173 | 0.232 | 0.280 | .379* | 0.335 | 0.270 | 0.328 | 0.291 | 0.331 | 0.274 | 0.242 | 0.265 | .361* | 0.277 | 0.221 | .360* | 0.277 | 0.239 |
| 2. Log Fp1 | 0.214 | 1.000 | .764** | .698** | .820** | .731** | .758** | .836** | .730** | .798** | .767** | .802** | .910** | .856** | .765** | .741** | .736** | .651** | .722** | .774** |
| 3. Log Fp2 | 0.173 | .764** | 1.000 | .672** | .760** | .739** | .783** | .771** | .838** | .843** | .684** | .702** | .752** | .813** | .746** | .719** | .723** | .781** | .856** | .878** |
| 4. Log F3 | 0.232 | .698** | .672** | 1.000 | .818** | .877** | .911** | .783** | .754** | .867** | .793** | .825** | .734** | .855** | .854** | .855** | .762** | .876** | .839** | .824** |
| 5. Log F4 | 0.280 | .820** | .760** | .818** | 1.000 | .830** | .851** | .830** | .851** | .829** | .815** | .764** | .865** | .876** | .864** | .705** | .900** | .748** | .734** | .826** |
| 6. Log C3 | .379* | .731** | .739** | .877** | .830** | 1.000 | .877** | .898** | .866** | .849** | .755** | .793** | .759** | .791** | .864** | .901** | .808** | .912** | .877** | .849** |
| 7. Log C4 | 0.335 | .758** | .783** | .911** | .851** | .877** | 1.000 | .788** | .850** | .941** | .833** | .904** | .771** | .918** | .931** | .878** | .797** | .906** | .887** | .861** |
| 8. Log P3 | 0.270 | .836** | .771** | .783** | .830** | .898** | .788** | 1.000 | .868** | .778** | .810** | .734** | .873** | .820** | .801** | .887** | .837** | .795** | .774** | .873** |
| 9. Log P4 | 0.328 | .730** | .838** | .754** | .851** | .866** | .850** | .868** | 1.000 | .827** | .766** | .782** | .762** | .854** | .857** | .800** | .893** | .869** | .818** | .877** |
| 10. Log O1 | 0.291 | .798** | .843** | .867** | .829** | .849** | .941** | .778** | .827** | 1.000 | .801** | .869** | .808** | .903** | .868** | .848** | .769** | .884** | .878** | .912** |
| 11. Log O2 | 0.331 | .767** | .684** | .793** | .815** | .755** | .833** | .810** | .766** | .801** | 1.000 | .816** | .818** | .862** | .836** | .739** | .775** | .728** | .716** | .851** |
| 12. Log F7 | 0.274 | .802** | .702** | .825** | .764** | .793** | .904** | .734** | .782** | .869** | .816** | 1.000 | .685** | .927** | .818** | .816** | .723** | .826** | .880** | .837** |
| 13. Log F8 | 0.242 | .910** | .752** | .734** | .865** | .759** | .771** | .873** | .762** | .808** | .818** | .685** | 1.000 | .812** | .822** | .722** | .807** | .661** | .636** | .803** |
| 14. Log T3 | 0.265 | .856** | .813** | .855** | .876** | .791** | .918** | .820** | .854** | .903** | .862** | .927** | .812** | 1.000 | .874** | .824** | .803** | .832** | .861** | .900** |
| 15. Log T4 | .361* | .765** | .746** | .854** | .864** | .864** | .931** | .801** | .857** | .868** | .836** | .818** | .822** | .874** | 1.000 | .817** | .825** | .822** | .801** | .787** |
| 16. Log T5 | 0.277 | .741** | .719** | .855** | .705** | .901** | .878** | .887** | .800** | .848** | .739** | .816** | .722** | .824** | .817** | 1.000 | .723** | .883** | .868** | .824** |
| 17. Log T6 | 0.221 | .736** | .723** | .762** | .900** | .808** | .797** | .837** | .893** | .769** | .775** | .723** | .807** | .803** | .825** | .723** | 1.000 | .775** | .710** | .815** |
| 18. Log Fz | .360* | .651** | .781** | .876** | .748** | .912** | .906** | .795** | .869** | .884** | .728** | .826** | .661** | .832** | .822** | .883** | .775** | 1.000 | .929** | .876** |
| 19. Log Cz | 0.277 | .722** | .856** | .839** | .734** | .877** | .887** | .774** | .818** | .878** | .716** | .880** | .636** | .861** | .801** | .868** | .710** | .929** | 1.000 | .884** |
| 20. Log Pz | 0.239 | .774** | .878** | .824** | .826** | .849** | .861** | .873** | .877** | .912** | .851** | .837** | .803** | .900** | .787** | .824** | .815** | .876** | .884** | 1.000 |

**Figure 1.:**
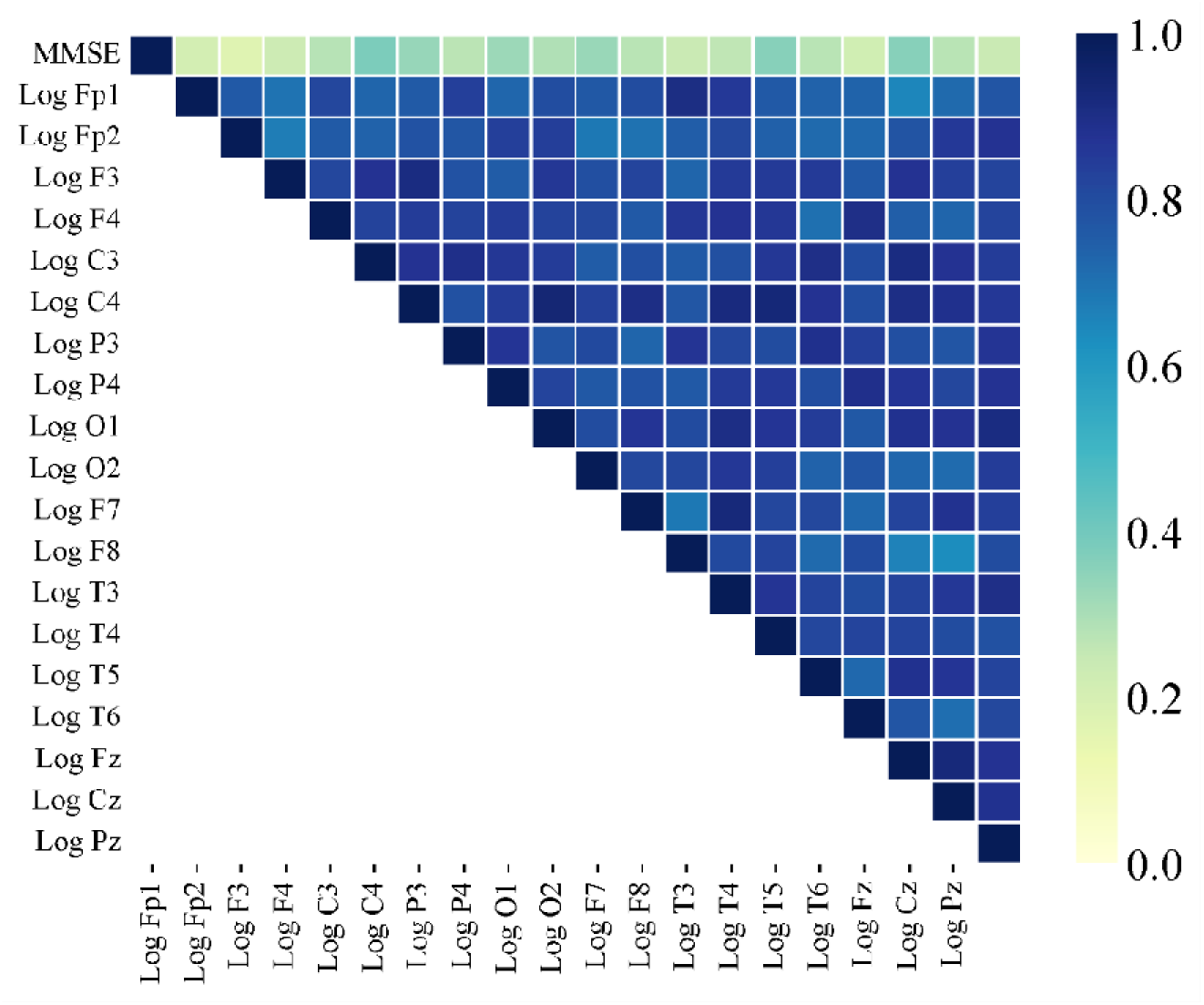
The heatmap reveals several variables with strong correlations to MMSE scores, a key measure of cognitive performance in AD. The strongest correlations are with Log Fp1, Log Fp2, Log Fp3, Log Fp4, Log F3, Log F4, Log F7, and Log F8, indicating that these EEG channels or frontal lobe regions are significant. Negative correlations, shown in dark blue, suggest that higher EEG values in these frontal regions are linked to worse cognitive performance or lower MMSE scores. This indicates that increased activity or abnormalities in the frontal lobe might signal cognitive decline in AD. In contrast, variables with weaker correlations to MMSE scores includ Log Cz, Log Pz, Log T5, Log T6, Log O1, and Log O2, which relate to central, parietal, and occipital lobes. This suggests that the frontal lobe is more crucial in cognitive performance and decline in AD patients.

For the FTD group, MMSE scores generally showed weak positive correlations with most EEG regions, but there were significant negative correlations with Log P3 (−0.234), Log P4 (−0.311), Log Cz (−0.376), and Log Pz (−0.344). These negative correlations suggest that higher EEG power in these specific region is linked to lower cognitive function. In other words, increased activity in Log P3, Log P4, Log Cz, and Log Pz corresponds to poorer performance on the MMSE, indicating that these regions are particularly important for understanding cognitive decline.

Additionally, the results revealed strong positive correlations among various EEG regions, indicating a coordinated pattern of brain activity. For example, Log Fp1 showed significant positive correlations with Log Fp2 (0.687**), Log F3 (0.594**), and Log C3 (0.637**), reflecting a high level of synchrony between these areas. Likewise, Log C3 and Log C4 demonstrated robust positive correlations with regions such as Log P3 (0.931**) and Log P4 (0.726**), suggesting a cohesive and interconnected network of EEG activity across these brain regions. The significant correlations are denoted by asterisks: **p < 0.01 and *p < 0.05. (table 3) (figure 2)

**Table 3.:** A correlation coefficient between various EEG electrodes and MMSE scores for individuals with FTD.

|  | 1 | 2 | 3 | 4 | 5 | 6 | 7 | 8 | 9 | 10 | 11 | 12 | 13 | 14 | 15 | 16 | 17 | 18 | 19 | 20 |
| --- | --- | --- | --- | --- | --- | --- | --- | --- | --- | --- | --- | --- | --- | --- | --- | --- | --- | --- | --- | --- |
| 1. MMSE | 1.000 | -0.030 | 0.039 | 0.250 | 0.057 | -0.041 | 0.054 | -0.234 | -0.311 | -0.014 | -0.138 | 0.109 | 0.071 | 0.051 | 0.052 | -0.054 | -0.028 | -0.064 | -0.376 | -0.344 |
| 2. Log Fp1 | -0.030 | 1.000 | .687** | .594** | .536* | .637** | .468* | .586** | .523* | .518* | .472* | .748** | .593** | .621** | .463* | .490* | .460* | .769** | .613** | .649** |
| 3. Log Fp2 | 0.039 | .687** | 1.000 | .529* | 0.423 | .458* | 0.309 | 0.395 | 0.377 | 0.307 | 0.278 | .460* | 0.363 | .487* | 0.210 | 0.360 | 0.347 | .650** | .491* | .448* |
| 4. Log F3 | 0.250 | .594** | .529* | 1.000 | .906** | .584** | .823** | .507* | .469* | .856** | .456* | .896** | .830** | .911** | .778** | .838** | .824** | .524* | 0.394 | .496* |
| 5. Log F4 | 0.057 | .536* | 0.423 | .906** | 1.000 | .553** | .813** | .607** | .551** | .850** | .435* | .885** | .870** | .868** | .784** | .815** | .785** | .532* | .517* | 0.413 |
| 6. Log C3 | -0.041 | .637** | .458* | .584** | .553** | 1.000 | .785** | .931** | .825** | .594** | .847** | .568** | .625** | .735** | .695** | .658** | .661** | .797** | .741** | .783** |
| 7. Log C4 | 0.054 | .468* | 0.309 | .823** | .813** | .785** | 1.000 | .775** | .726** | .835** | .626** | .753** | .864** | .942** | .929** | .905** | .901** | .499* | .550** | .578** |
| 8. Log P3 | -0.234 | .586** | 0.395 | .507* | .607** | .931** | .775** | 1.000 | .912** | .626** | .818** | .518* | .634** | .714** | .686** | .685** | .682** | .750** | .841** | .733** |
| 9. Log P4 | -0.311 | .523* | 0.377 | .469* | .551** | .825** | .726** | .912** | 1.000 | .654** | .774** | .474* | .523* | .708** | .609** | .753** | .742** | .639** | .718** | .716** |
| 10. Log O1 | -0.014 | .518* | 0.307 | .856** | .850** | .594** | .835** | .626** | .654** | 1.000 | .638** | .820** | .778** | .872** | .743** | .921** | .904** | 0.388 | 0.421 | .570** |
| 11. Log O2 | -0.138 | .472* | 0.278 | .456* | .435* | .847** | .626** | .818** | .774** | .638** | 1.000 | 0.422 | .445* | .592** | .473* | .642** | .621** | .675** | .573** | .720** |
| 12. Log F7 | 0.109 | .748** | .460* | .896** | .885** | .568** | .753** | .518* | .474* | .820** | 0.422 | 1.000 | .864** | .849** | .749** | .748** | .711** | .549** | 0.424 | .496* |
| 13. Log F8 | 0.071 | .593** | 0.363 | .830** | .870** | .625** | .864** | .634** | .523* | .778** | .445* | .864** | 1.000 | .887** | .881** | .781** | .794** | .546* | .596** | .471* |
| 14. Log T3 | 0.051 | .621** | .487* | .911** | .868** | .735** | .942** | .714** | .708** | .872** | .592** | .849** | .887** | 1.000 | .877** | .941** | .934** | .606** | .581** | .633** |
| 15. Log T4 | 0.052 | .463* | 0.210 | .778** | .784** | .695** | .929** | .686** | .609** | .743** | .473* | .749** | .881** | .877** | 1.000 | .825** | .841** | .479* | .527* | .509* |
| 16. Log T5 | -0.054 | .490* | 0.360 | .838** | .815** | .658** | .905** | .685** | .753** | .921** | .642** | .748** | .781** | .941** | .825** | 1.000 | .984** | .484* | .485* | .610** |
| 17. Log T6 | -0.028 | .460* | 0.347 | .824** | .785** | .661** | .901** | .682** | .742** | .904** | .621** | .711** | .794** | .934** | .841** | .984** | 1.000 | .468* | .516* | .627** |
| 18. Log Fz | -0.064 | .769** | .650** | .524* | .532* | .797** | .499* | .750** | .639** | 0.388 | .675** | .549** | .546* | .606** | .479* | .484* | .468* | 1.000 | .778** | .674** |
| 19. Log Cz | -0.376 | .613** | .491* | 0.394 | .517* | .741** | .550** | .841** | .718** | 0.421 | .573** | 0.424 | .596** | .581** | .527* | .485* | .516* | .778** | 1.000 | .765** |
| 20. Log Pz | -0.344 | .649** | .448* | .496* | 0.413 | .783** | .578** | .733** | .716** | .570** | .720** | .496* | .471* | .633** | .509* | .610** | .627** | .674** | .765** | 1.000 |

**Figure 2.:**
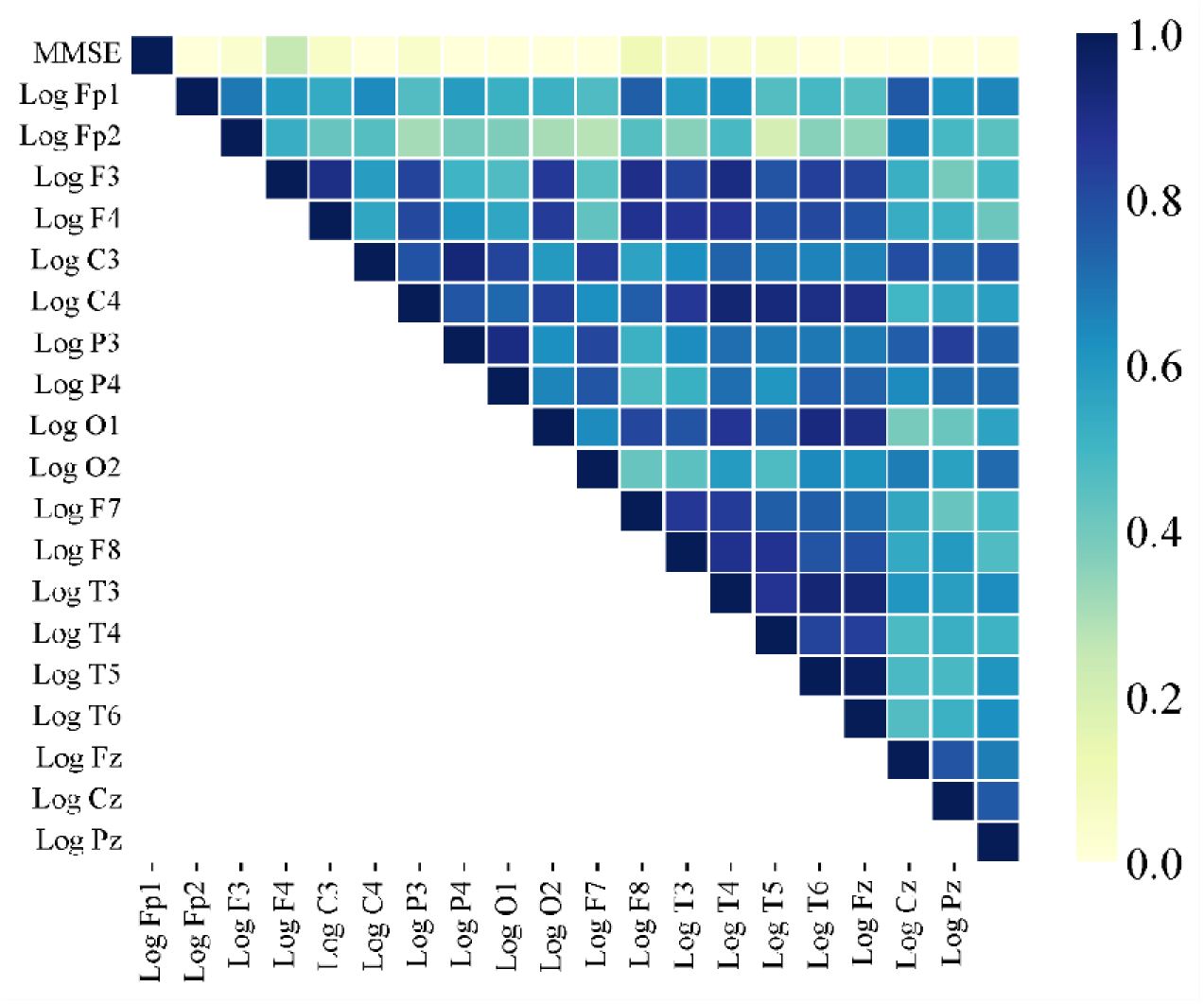
The figure is a correlation matrix heatmap illustrating the relationships between MMSE scores an various logarithmic EEG measures from different brain regions in FTD patients. The top row shows that the strongest correlations with MMSE scores are found in the frontal lobe regions (Log Fp1, Log Fp2, Log F3, Log F4, Log F7, Log F8), indicated by lighter colors, suggesting these areas are critical for cognitive performance. I contrast, weaker correlations are observed in central, parietal, and occipital regions (Log C3, Log C4, Log P3, Log P4, Log O1, Log O2, Log T3, Log T4, Log T5, Log T6, Log Fz, Log Cz, Log Pz), shown by darker colors, indicating these areas are less influential in cognitive decline as measured by the MMSE. High inter-correlations within the frontal lobe regions further emphasize their importance in cognitive function, potentially serving as biomarkers for assessing cognitive health in FTD patients.

Using linear regression, we examined the relationship between EEG data from various brain channels and cognitive scores (MMSE), while accounting for age and gender. This analysis aimed to identify whether specific EEG patterns are associated with cognitive decline and how well these patterns predict changes in cognitive performance. (Table 4)

**Table 4.:** Linear Regression Analysis of EEG Channels and MMSE Scores, Age, and Gender.

| AD |  |  |  |  |  |  | FTD |  |  |  |  |  |
| --- | --- | --- | --- | --- | --- | --- | --- | --- | --- | --- | --- | --- |
| Model | Adjusted R<sup>2</sup> | B (S.E.) | $\beta$ | 95% CI | | P value | Adjusted R<sup>2</sup> | B (S.E.) | $\beta$ | 95% CI | | P value |
|  |  |  |  | lower | Upper |  |  |  |  | lower | Upper |  |
| Log Fp1 | 0.025 |  |  |  |  |  | 0.012 |  |  |  |  |  |
| MMSE |  | 0.118 (0.095) | 1.241 | -0.076 | 0.312 | 0.223 |  | -0.028 (0.21) | -0.133 | -0.467 | 0.412 | 0.896 |
| Gender |  | -0.784 (0.87) | -0.9 | -2.557 | 0.99 | 0.375 |  | -0.285 (1.11) | -0.257 | -2.61 | 2.039 | 0.8 |
| Age |  | -0.074 (0.054) | -1.363 | -0.184 | 0.036 | 0.182 |  | 0.103 (0.06) | 1.732 | -0.22 | 0.228 | 0.099 |
| Log Fp2 | -0.028 |  |  |  |  |  | -0.032 |  |  |  |  |  |
| MMSE |  | 0.098 (0.098) | 0.994 | -0.102 | 0.297 | 0.328 |  | 0.034 (0.202) | 0.169 | -0.389 | 0.458 | 0.868 |
| Gender |  | -0.751 (0.897) | -0.838 | -2.578 | 1.075 | 0.408 |  | -0.917 (1.07) | -0.857 | -3.156 | 1.323 | 0.402 |
| Age |  | -0.032 (0.056) | 0.58 | -0.146 | 0.081 | 0.556 |  | -0.068 (0.058) | 1.173 | -0.053 | 0.188 | 0.255 |
| Log F3 | -0.034 |  |  |  |  |  | -0.07 |  |  |  |  |  |
| MMSE |  | 0.133 (0.098) | 1.351 | -0.067 | 0.333 | 0.186 |  | 0.253 (0.224) | 1.126 | -0.217 | 0.732 | 0.274 |
| Gender |  | 0.052 (0.899) | 0.058 | -1.778 | 1.883 | 0.954 |  | -1.127 (1.187) | -0.949 | -3.612 | 1.358 | 0.354 |
| Age |  | -0.026 (0.056) | -0.471 | -0.14 | 0.088 | 0.641 |  | -0.014 (0.064) | -0.22 | -0.148 | 0.12 | 0.828 |
| Log F4 | 0.044 |  |  |  |  |  | -0.097 |  |  |  |  |  |
| MMSE |  | 0.155 (0.094) | 1.653 | -0.036 | 0.347 | 0.108 |  | 0.058 (0.233) | 0.248 | -0.429 | 0.545 | 0.806 |
| Gender |  | -0.58 (0.859) | -0.676 | -2.329 | 1.169 | 0.504 |  | -1.122 (1.231) | -0.911 | -3.699 | 1.456 | 0.374 |
| Age |  | -0.075 (0.053) | -1.409 | -0.184 | 0.034 | 0.169 |  | -0.032 (0.066) | -0.479 | -0.17 | 0.107 | 0.638 |
| Log C3 | 0.089 |  |  |  |  |  | 0.183 |  |  |  |  |  |
| MMSE |  | 0.213 (0.0920) | 2.319 | 0.026 | 0.4 | **0.027** |  | -0.031 (0.173) | -0.18 | -0.394 | 0.331 | 0.859 |
| Gender |  | -0.525 (0.841) | -0.624 | -2.237 | 1.188 | 0.537 |  | -0.704 (0.916) | -0.768 | -2.621 | 1.213 | 0.452 |
| Age |  | -0.054 (0.052) | -1.029 | -0.16 | 0.053 | 0.311 |  | 0.126 (0.049) | 2.559 | 0.023 | 0.229 | **0.019** |
| Log C4 | 0.118 |  |  |  |  |  | -0.1 |  |  |  |  |  |
| MMSE |  | 0.19 (0.095) | 2.012 | -0.002 | 0.383 | 0.053 |  | 0.055 (0.233) | 0.238 | -0.432 | 0.543 | 0.815 |
| Gender |  | -0.131 (0.865) | -0.151 | -1.893 | 1.632 | 0.881 |  | -0.704 (1.231) | -0.572 | -3.281 | 1.873 | 0.574 |
| Age |  | -0.042 (0.054) | -0.787 | -0.152 | 0.067 | 0.437 |  | 0.052 (0.066) | 0.788 | -0.086 | 0.191 | 0.441 |
| Log P3 | 0.086 |  |  |  |  |  | 0.236 |  |  |  |  |  |
| MMSE |  | 0.149 (0.094) | 1.586 | -0.042 | 0.34 | 0.123 |  | -0.179 (0.17) | -1.051 | -0.536 | 0.178 | 0.306 |
| Gender |  | -1.331 (0.858) | -1.551 | -3.079 | 0.418 | 0.131 |  | -0.725 (0.902) | -0.804 | -2.612 | 1.162 | 0.431 |
| Age |  | -0.064 (0.053) | -1.194 | -0.172 | 0.045 | 0.241 |  | 0.117 (0.048) | 2.408 | 0.015 | 0.218 | **0.026** |
| Log P4 | 0.073 |  |  |  |  |  | 0.135 |  |  |  |  |  |
| MMSE |  | 0.186 (0.094) | 1.966 | -0.007 | 0.378 | 0.058 |  | -0.273 (0.192) | -1.424 | -0.674 | 0.128 | 0.171 |
| Gender |  | -0.881 (0.964) | -1.02 | -2.64 | 0.878 | 0.315 |  | 0.068 (1.013) | 0.067 | -2.052 | 2.189 | 0.947 |
| Age |  | -0.055 (0.054) | -1.022 | -0.164 | 0.055 | 0.314 |  | 0.102 (0.054) | 1.876 | -0.012 | 0.216 | 0.076 |
| Log O1 | 0.008 |  |  |  |  |  | -0.154 |  |  |  |  |  |
| MMSE |  | 0.164 (0.095) | 1.724 | -0.03 | 0.358 | 0.094 |  | -0.015 (0.241) | -0.063 | -0.519 | 0.489 | 0.951 |
| Gender |  | -0.229 (0.871) | -0.263 | -2.002 | 1.545 | 0.794 |  | 0.253 (1.273) | 0.199 | -2.411 | 2.918 | 0.845 |
| Age |  | -0.04 (0.054) | -0.744 | -0.151 | 0.07 | 0.462 |  | 0.013 (0.068) | 0.184 | -0.131 | 0.156 | 0.856 |
| Log O2 | 0.139 |  |  |  |  |  | 0.095 |  |  |  |  |  |
| MMSE |  | 0.181 (0.091) | 1.983 | -0.005 | 0.367 | 0.056 |  | -0.113 (0.187) | -0.606 | -0.504 | 0.278 | 0.552 |
| Gender |  | -1.128 (0.835) | -1.351 | -2.828 | 0.572 | 0.186 |  | 0.462 (0.988) | 0.467 | -1.606 | 2.53 | 0.646 |
| Age |  | -0.098 (0.052) | -1.883 | -0.203 | 0.008 | 0.069 |  | 0.116 (0.053) | 2.191 | 0.005 | 0.228 | **0.041** |
| Log F7 | 0.001 |  |  |  |  |  | -0.144 |  |  |  |  |  |
| MMSE |  | 0.165 (0.102) | 1.614 | -0.043 | 0.373 | 0.116 |  | 0.118 (0.246) | 0.48 | -0.397 | 0.633 | 0.637 |
| Gender |  | -0.234 (0.933) | -0.251 | -2.135 | 1.666 | 0.803 |  | -0.28 (1.301) | -0.215 | -3.004 | 2.443 | 0.832 |
| Age |  | -0.051 (0.058) | -0.881 | -0.169 | 0.067 | 0.385 |  | -0.001 (0.07) | -0.015 | -0.148 | 0.145 | 0.988 |
| Log F8 | 0.101 |  |  |  |  |  | -0.112 |  |  |  |  |  |
| MMSE |  | 0.136 (0.097) | 1.411 | -0.061 | 0.333 | 0.168 |  | 0.075 (0.242) | 0.308 | -0.433 | 0.582 | 0.761 |
| Gender |  | -0.793 (0.884) | -0.898 | -2.594 | 1.007 | 0.376 |  | -1.089 (1.28) | -0.85 | -3.772 | 1.594 | 0.406 |
| Age |  | -0.121 (0.055) | -2.206 | -0.233 | -0.009 | **0.035** |  | -0.019 (0.069) | -0.278 | -0.163 | 0.125 | 0.784 |
| Log T3 | 0.02 |  |  |  |  |  | -0.116 |  |  |  |  |  |
| MMSE |  | 0.151 (0.097) | 1.552 | -0.047 | 0.348 | 0.13 |  | 0.052 (0.231) | 0.224 | -0.432 | 0.536 | 0.825 |
| Gender |  | -0.718 (0.887) | -0.809 | -2.525 | 1.09 | 0.425 |  | -1.011 (1.223) | -0.827 | -3.571 | 1.549 | 0.419 |
| Age |  | -0.05 90.056) | -0.912 | -0.163 | 0.062 | 0.369 |  | -0.002 (0.066) | -0.027 | -0.139 | 0.136 | 0.979 |
| Log T4 | 0.072 |  |  |  |  |  | -0.144 |  |  |  |  |  |
| MMSE |  | 0.211 (0.096) | 2.19 | 0.015 | 0.406 | **0.036** |  | 0.054 (0.242) | 0.225 | -0.452 | 0.56 | 0.825 |
| Gender |  | 0.06 (0.879) | 0.068 | -1.731 | 1.85 | 0.946 |  | 0.242 (1.278) | 0.19 | -2.433 | 2.918 | 0.852 |
| Age |  | -0.79 (0.055) | -1.441 | -0.19 | 0.033 | 0.159 |  | 0.021 (0.069) | 0.301 | -0.123 | 0.165 | 0.767 |
| Log T5 | 0.027 |  |  |  |  |  | -0.142 |  |  |  |  |  |
| MMSE |  | 0.156 (0.096) | 1.629 | -0.039 | 0.351 | 0.113 |  | -0.059 (0.25) | -0.237 | -0.582 | 0.464 | 0.815 |
| Gender |  | -0.852 (0.875) | -0.974 | -2.635 | 0.93 | 0.337 |  | -0.374 (1.321) | -0.238 | -3.139 | 2.392 | 0.78 |
| Age |  | -0.21 (0.054) | -0.386 | -0.132 | 0.09 | 0.702 |  | -0.009 (0.071) | -0.132 | -0.158 | 0.139 | 0.896 |
| Log T6 | 0.067 |  |  |  |  |  | -0.131 |  |  |  |  |  |
| MMSE |  | 0.117 (0.091) | 1.285 | -0.069 | 0.303 | 0.208 |  | -0.031 (0.255) | -0.122 | -0.565 | 0.502 | 0.904 |
| Gender |  | -1.031 (0.834) | -1.236 | -2.73 | 0.669 | 0.226 |  | -0.645 (-0.479) | -0.479 | -3.468 | 2.177 | 0.638 |
| Age |  | -0.084 (0.052) | -1.619 | -0.19 | 0.022 | 0.115 |  | -0.023 (0.073) | -0.318 | -0.175 | 0.129 | 0.754 |
| Log Fz | 0.096 |  |  |  |  |  | 0.059 |  |  |  |  |  |
| MMSE |  | 0.2 (0.092) | 2.18 | 0.013 | 0.387 | **0.037** |  | -0.05 (0.1810) | -0.278 | -0.429 | 0.329 | 0.784 |
| Gender |  | -0.934 (0.839) | -1.113 | -2.644 | 0.776 | 0.274 |  | -1.237 (0.958) | -1.291 | -3.243 | 0.768 | 0.212 |
| Age |  | -0.045 (0.052) | -0.863 | -0.151 | 0.061 | 0.394 |  | 0.061 (0.052) | 1.189 | -0.047 | 0.169 | 0.249 |
| Log Cz | 0.006 |  |  |  |  |  | 0.273 |  |  |  |  |  |
| MMSE |  | 0.157 (0.096) | 1.628 | -0.039 | 0.353 | 0.113 |  | -0.299 (0.169) | -1.769 | -0.652 | 0.055 | 0.093 |
| Gender |  | -0.458 (0.88) | -0.52 | -2.251 | 1.335 | 0.606 |  | -1.073 (0.892) | -1.203 | -2.941 | 0.795 | 0.244 |
| Age |  | -0.005 (0.055) | -0.097 | -0.117 | 0.106 | 0.923 |  | 0.07 (0.048) | 1.461 | -0.03 | 0.171 | 0.16 |
| Log Pz | 0.041 |  |  |  |  |  | 0.183 |  |  |  |  |  |
| MMSE |  | 0.131 (0.094) | 1.393 | -0.061 | 0.323 | 0.173 |  | -0.288 (0.180) | -1.599 | -0.666 | 0.089 | 0.126 |
| Gender |  | -1.069 (0.86) | -1.244 | -2.821 | 0.682 | 0.223 |  | -0.268 (0.954) | -0.281 | -2.264 | 1.728 | 0.782 |
| Age |  | -0.056 (0.053) | -1.049 | -0.165 | 0.053 | 0.302 |  | 0.095 (0.051) | 1.85 | -0.012 | 0.202 | 0.08 |

In AD, the EEG data analysis revealed that most channels do not show significant correlations with MMSE scores, indicating that cognitive decline as measured by MMSE may not strongly influence EEG patterns in this dataset. Notably, channels like Log C3 (B=2.319 and p=0.027), Log C4 (B=2.012 and p=0.053), Log T4 (B = 2.19, p = 0.036), and Log Fz (B = 2.18, p = 0.037) showed marginally significant or significant associations with MMSE scores. Besides, Log C3 (B = 2.319, p = 0.027) showed a significant positive correlation, suggesting that lower MMSE scores (indicating worse cognitive performance) are associated with increased activity in this channel. However, age had mixed effects, with only a few channels like Log F8 (B = −2.206, p = 0.035) showing significant negative correlations, implying that age-related changes in EEG patterns are not uniform across all channels.

In FTD, there was a consistent and significant negative relationship between age and EEG activity across most channels. This suggests that as patients age, there is a marked decline in EEG activity, reflecting the progressive nature of the disease. Channels such as Log Fp1, Log Fp2, Log C3, and Log O2 showed significant negative correlations with age, meaning that older age is associated with reduced EEG activity in these regions. This widespread negative association underscores how aging can impact EEG patterns in FTD patients, potentially reflecting disease-related alterations in brain function.

## 4. Discussion

Our study highlighted the distinct relationships between cognitive function, age, and brain activity in AD and FTD. We found that cognitive function using MMSE has a more significant relationship with brain activity in AD, whereas age has a more prominent impact on brain activity in FTD.

As per findings, in AD, the MMSE was a significant predictor of brain activity in regions such as C3, C4, T4, and Fz, indicating a close relationship between cognitive function and activity in these areas. Correlation analysis supported this, showing significant positive associations between MMSE scores and EEG power in Log Fp1, Log T4, and Log Fz, suggesting that better cognitive performance is related to higher EEG power in frontal and temporal regions. Additionally, age was a significant predictor of brain activity in the F8 region, indicating age-related changes in brain activity.

Nonetheless, in FTD, age was a significant predictor of brain activity in regions such as C3, P3, O1, and O2, indicating a more consistent and widespread impact compared to AD. MMSE was not a significant predictor in any region for FTD, suggesting that cognitive function is less related to brain activity in FTD patients. Correlation analysis showed significant negative correlations between MMSE scores and EEG power in Log P3, Log P4, Log Cz, and Log Pz, indicating that higher EEG power in these regions is linked to lower cognitive function in FTD patients.

Our findings align with previous research indicating that cognitive decline in AD is associated with specific changes in brain activity, particularly in the frontal and temporal regions [42–47]. Cognitive decline in AD is characterized by a progressive deterioration of cognitive functions, which can be measured using techniques such as EEG. Research has consistently shown that the frontal and temporal regions of the brain are crucial for various cognitive functions [48]. For instance, the frontal regions are involved in executive functions, such as planning, decision-making, problem-solving, and controlling behavior [49, 50], while the temporal regions play a significant role in memory and language processing, containing structures like the hippocampus, which is critical for forming and retrieving memories [51–53].

In AD, the frontal and temporal regions are among the first to show significant pathological changes, including the accumulation of amyloid plaques and neurofibrillary tangles. These pathological changes lead to neuron damage and death, resulting in impairments in memory, executive function, and language. Patients struggle with forming new memories and recalling past events, face difficulties with tasks requiring planning and organization, and experience challenges with word finding and understanding language [54–56].

Our study found significant positive correlations between MMSE scores and EEG power in regions such as Fp1 (frontal pole 1), T4 (right temporal lobe), and Fz (midline frontal). These findings reinforce the understanding that specific changes in brain activity, detectable through EEG, are related to cognitive decline in AD. Researchers, by identifying these correlations, can better understand the neural mechanisms underlying cognitive decline and potentially develop targeted interventions to slow or mitigate the effects of AD.

Another study found that Neurometric QEEG measures, particularly increased theta power and delta power in later stages, are sensitive indicators of cognitive impairment, especially significant in a bilateral temporo-parietal arc, suggesting their potential utility in the early evaluation of dementia and estimation of cognitive deterioration in Alzheimer’s type dementia patients [57]. Likewise, cognitive impairment in patients with epilepsy (PWE) was significantly associated with QEEG measures, particularly interhemispheric delta and beta coherences, suggesting that QEEG may be useful in understanding the pathophysiological basis of cognitive alterations in epilepsy [58]. Notably, a former study found that there are significant gender differences in brain organization and functioning, with women displaying higher interhemispheric synchronization and lower hemispheric differentiation than men. This means that, in women, higher interhemispheric correlation was positively associated with abstract and spatial aptitude scores in the central cortex, whereas in men, it was negatively associated with spatial, abstract, and verbal aptitude scores across most recorded brain regions [59].

Our results in FTD align with earlier studies that indicate a broader impact of age on brain activity [24, 60]. Unlike AD, where specific brain regions were primarily affected, FTD showed a more diffuse pattern of brain activity changes. This broader impact may reflect the different underlying mechanisms of FTD compared to AD, where the disease pathology is not confined to specific areas but rather spreads more widely across the brain.

In FTD, we did not observe a significant relationship between MMSE scores and brain activity. This finding is consistent with the understanding that FTD affects the brain more diffusely, disrupting cognitive functions in a less region-specific manner. The MMSE, while useful in assessing general cognitive function, may not be as effective in capturing the complex and widespread brain changes occurring in FTD. Interestingly, we found negative correlations between MMSE scores and EEG power in the parietal regions (P3, P4, Cz, Pz). These regions are involved in various cognitive processes, including spatial orientation, sensory perception, and integration of sensory information. The negative correlations suggest that higher cognitive impairment in FTD is associated with increased EEG power in these areas. This could indicate compensatory neural activity or pathological overactivation in response to neurodegenerative changes.

Aging contributes to the development and progression of dementia through a combination of cellular and molecular mechanisms [24, 61, 62]. At the cellular level, increased oxidative stress [63–65], mitochondrial dysfunction [66, 67], and impaired autophagy [68] lead to the accumulation of damaged proteins and organelles, causing neuronal damage. Chronic inflammation and immune response dysregulation further exacerbate neurodegeneration [68–70]. Thus, understanding these mechanisms provides insights into the complex relationship between aging and dementia, potentially guiding the development of targeted therapies to mitigate age-related cognitive decline.

The implications of the current study are twofold. First, it emphasizes the importance of specific brain regions in cognitive performance, suggesting potential targets for neurostimulation or other therapeutic interventions aimed at enhancing cognitive function. Second, it underscores the need for a nuanced approach in analyzing EEG data, recognizing that not all brain regions equally contribute to cognitive abilities as measured by global scales like the MMSE.

Future studies should consider several key limitations and confounders when investigating the relationship between EEG activity and cognitive function, as observed in this study. These include potential constraints related to sample size and generalizability, given the need for larger and more diverse participant cohorts to ensure robust findings across different populations. Notice that addressing selection biases in participant recruitment and standardizing electrode placement and signal interpretation methods are crucial to minimize variability in EEG data. Moreover, considering the influence of medications, comorbidities, and participant characteristics on EEG readings and cognitive outcomes is essential for accurate interpretation. Methodologically, future research could benefit from integrating multimodal imaging techniques such as fMRI or PET and advanced statistical analyses to explore network dynamics and uncover nuanced associations between brain regions and specific cognitive domains. Thus, by addressing these factors, future studies can enhance the reliability and applicability of findings in understanding the neural underpinnings of cognitive function across various neurological conditions.

## 5. Conclusion

This study aimed to investigate regional differences in EEG patterns among AD and FTD cases to enhance diagnostic strategies. It highlighted distinct relationships between cognitive function, age, and brain activity in AD and FTD. In AD, cognitive function measured by MMSE scores significantly predicted brain activity, particularly in the frontal and temporal regions, indicating a close relationship between cognitive performance and EEG power in these areas. Conversely, in FTD, age was a more significant predictor of brain activity, with a consistent and widespread impact across multiple brain regions. The findings suggest that EEG biomarkers can enhance diagnostic strategies by highlighting different patterns of brain activity related to cognitive function and age in AD and FTD.

## Abbreviations

EEG: electroencephalography
AD: Alzheimer’s disease
FTD: frontotemporal dementia
MMSE: Mini-Mental State Examination.

## Declarations section

## Acknowledgments

We acknowledge data descriptor (https://doi.org/10.3390/data8060095) and OpenNeuro that provided this data.

## Ethical Approval and Consent to Participate

The study was conducted in accordance with the Declaration of Helsinki and approved by the Scientific and Ethics Committee of AHEPA University Hospital, Aristotle University of Thessaloniki, under protocol number 142/12-04-2023.

## Consent for Publication

Not applicable.

## Authors’ Contributions

All authors listed have made a substantial, direct, and intellectual contribution to the work, and approved it for publication. A.A and H.N. contributed to developing research ideas and analysis. A.A., H.N., and F.A. contributed to interpretation of data, writing the draft, and revising it. All authors read and approved the final manuscript.

## Availability of Data and Materials

The data for this article were obtained from OpenNeuro Dataset ds004504. Dataset DOI is https://doi.org/10.18112/openneuro.ds004504.v1.0.7.

## References

1. Popov T, Tröndle M, Baranczuk-Turska Z, Pfeiffer C, Haufe S, Langer N. Test–retest reliability of resting-state EEG in young and older adults. Psychophysiology. 2023;60(7):e14268.

2. Da Silva FL. EEG: origin and measurement. EEG-fMRI: physiological basis, technique, and applications. Springer; 2023. p. 23–48.

3. Perez V, Duque A, Hidalgo V, Salvador A. EEG Frequency Bands in Subjective Cognitive Decline: A Systematic Review of Resting State Studies. Biological Psychology. 2024:108823.

4. Ward D, Jones R, Bones PJ, Carroll G. Enhancement of deep epileptiform activity in the EEG via 3-D adaptive spatial filtering. IEEE transactions on bio-medical engineering. 1999;46:707–16. doi: 10.1109/10.764947.

5. Babiloni C, Triggiani AI, Lizio R, Cordone S, Tattoli G, Bevilacqua V, et al. Classification of Single Normal and Alzheimer’s Disease Individuals from Cortical Sources of Resting State EEG Rhythms. Frontiers in Neuroscience. 2016;10. doi: 10.3389/fnins.2016.00047.

6. Kim S-E, Shin C, Yim J, Seo K, Ryu H, Choi H, et al. Resting-state electroencephalographic characteristics related to mild cognitive impairments. Frontiers in Psychiatry. 2023;14. doi: 10.3389/fpsyt.2023.1231861.

7. Zheng X, Wang B, Liu H, Wu W, Sun J, Fang W, et al. Diagnosis of Alzheimer’s disease via resting-state EEG: integration of spectrum, complexity, and synchronization signal features. Frontiers in Aging Neuroscience. 2023;15. doi: 10.3389/fnagi.2023.1288295.

8. Cassani R, Estarellas M, San-Martin R, Fraga FJ, Falk TH. Systematic Review on Resting-State EEG for Alzheimer’s Disease Diagnosis and Progression Assessment. Dis Markers. 2018;2018:5174815. doi: 10.1155/2018/5174815.

9. Aoki Y, Takahashi R, Suzuki Y, Pascual-Marqui RD, Kito Y, Hikida S, et al. EEG resting-state networks in Alzheimer’s disease associated with clinical symptoms. Scientific Reports. 2023;13(1):3964. doi: 10.1038/s41598-023-30075-3.

10. Kopčanová M, Tait L, Donoghue T, Stothart G, Smith L, Flores-Sandoval AA, et al. Resting-state EEG signatures of Alzheimer’s disease are driven by periodic but not aperiodic changes. Neurobiol Dis. 2024;190:106380. doi: 10.1016/j.nbd.2023.106380.

11. Nardone R, Sebastianelli L, Versace V, Saltuari L, Lochner P, Frey V, et al. Usefulness of EEG Techniques in Distinguishing Frontotemporal Dementia from Alzheimer’s Disease and Other Dementias. Dis Markers. 2018;2018:6581490. doi: 10.1155/2018/6581490.

12. Lal U, Chikkankod AV, Longo L. A Comparative Study on Feature Extraction Techniques for the Discrimination of Frontotemporal Dementia and Alzheimer’s Disease with Electroencephalography in Resting-State Adults. Brain Sciences. 2024 doi: 10.3390/brainsci14040335.

13. Si Y, He R, Jiang L, Yao D, Zhang H, Xu P, et al. Differentiating Between Alzheimer’s Disease and Frontotemporal Dementia Based on the Resting-State Multilayer EEG Network. IEEE Transactions on Neural Systems and Rehabilitation Engineering. 2023;31:4521–7. doi: 10.1109/TNSRE.2023.3329174.

14. Moretti DV, paternicò d, Binetti G, Zanetti O, Frisoni gb. EEG upper/low alpha frequency power ratio relates to temporo-parietal brain atrophy and memory performances in mild cognitive impairment. Frontiers in Aging Neuroscience. 2013;5. doi: 10.3389/fnagi.2013.00063.

15. Lejko N, Larabi DI, Herrmann CS, Aleman A, Ćurčić-Blake B. Alpha Power and Functional Connectivity in Cognitive Decline: A Systematic Review and Meta-Analysis. J Alzheimers Dis. 2020;78(3):1047-88. doi: 10.3233/jad-200962.

16. Jiao B, Li R, Zhou H, Qing K, Liu H, Pan H, et al. Neural biomarker diagnosis and prediction to mild cognitive impairment and Alzheimer’s disease using EEG technology. Alzheimer’s Research & Therapy. 2023;15(1):32. doi: 10.1186/s13195-023-01181-1.

17. Arevalo-Rodriguez I, Smailagic N, Roqué-Figuls M, Ciapponi A, Sanchez-Perez E, Giannakou A, et al. Mini-Mental State Examination (MMSE) for the early detection of dementia in people with mild cognitive impairment (MCI). Cochrane Database Syst Rev. 2021;7(7):Cd010783. doi: 10.1002/14651858.CD010783.pub3.

18. Arevalo-Rodriguez I, Smailagic N, Roqué IFM, Ciapponi A, Sanchez-Perez E, Giannakou A, et al. Mini-Mental State Examination (MMSE) for the detection of Alzheimer’s disease and other dementias in people with mild cognitive impairment (MCI). Cochrane Database Syst Rev. 2015;2015(3):Cd010783. doi: 10.1002/14651858.CD010783.pub2.

19. Torabinikjeh M, Asayesh V, Dehghani M, Kouchakzadeh A, Marhamati H, Gharibzadeh S. Correlations of frontal resting-state EEG markers with MMSE scores in patients with Alzheimer’s disease. The Egyptian Journal of Neurology, Psychiatry and Neurosurgery. 2022;58(1):31. doi: 10.1186/s41983-022-00465-x.

20. Lee SJ, Park MH, Park SS, Ahn JY, Heo JH. Quantitative EEG and medial temporal lobe atrophy in Alzheimer’s dementia: Preliminary study. Ann Indian Acad Neurol. 2015;18(1):10–4. doi: 10.4103/0972-2327.145284.

21. Xu M, Zhang Y, Zhang Y, Liu X, Qing K. EEG biomarkers analysis in different cognitive impairment after stroke: an exploration study. Front Neurol. 2024;15:1358167. doi: 10.3389/fneur.2024.1358167.

22. Araújo T, Teixeira J, Rodrigues P. Smart-Data-Driven System for Alzheimer Disease Detection through Electroencephalographic Signals. Bioengineering. 2022;9:141. doi: 10.3390/bioengineering9040141.

23. Caravaglios G, Muscoso EG, Blandino V, Di Maria G, Gangitano M, Graziano F, et al. EEG Resting-State Functional Networks in Amnestic Mild Cognitive Impairment. Clinical EEG and Neuroscience. 2022;54(1):36–50. doi: 10.1177/15500594221110036.

24. Lee J, Kim H-J. Normal Aging Induces Changes in the Brain and Neurodegeneration Progress: Review of the Structural, Biochemical, Metabolic, Cellular, and Molecular Changes. Frontiers in Aging Neuroscience. 2022;14. doi: 10.3389/fnagi.2022.931536.

25. Pavlov AN, Pitsik EN, Frolov NS, Badarin A, Pavlova ON, Hramov AE. Age-Related Distinctions in EEG Signals during Execution of Motor Tasks Characterized in Terms of Long-Range Correlations. Sensors (Basel). 2020;20(20). doi: 10.3390/s20205843.

26. Javaid H, Kumarnsit E, Chatpun S. Age-Related Alterations in EEG Network Connectivity in Healthy Aging. Brain Sci. 2022;12(2). doi: 10.3390/brainsci12020218.

27. He M, Liu F, Nummenmaa A, Hämäläinen M, Dickerson BC, Purdon PL. Age-Related EEG Power Reductions Cannot Be Explained by Changes of the Conductivity Distribution in the Head Due to Brain Atrophy. Frontiers in Aging Neuroscience. 2021;13. doi: 10.3389/fnagi.2021.632310.

28. Hou Y, Dan X, Babbar M, Wei Y, Hasselbalch SG, Croteau DL, Bohr VA. Ageing as a risk factor for neurodegenerative disease. Nat Rev Neurol. 2019;15(10):565–81. doi: 10.1038/s41582-019-0244-7.

29. Kesidou E, Theotokis P, Damianidou O, Boziki M, Konstantinidou N, Taloumtzis C, et al. CNS Ageing in Health and Neurodegenerative Disorders. J Clin Med. 2023;12(6). doi: 10.3390/jcm12062255.

30. Kopčanová M, Tait L, Donoghue T, Stothart G, Smith L, Sandoval AAF, et al. Resting-state EEG signatures of Alzheimer’s disease are driven by periodic but not aperiodic changes. bioRxiv. 2023. doi: 10.1101/2023.06.11.544491.

31. Merkin A, Sghirripa S, Graetz L, Smith AE, Hordacre B, Harris R, et al. Do age-related differences in aperiodic neural activity explain differences in resting EEG alpha? Neurobiology of Aging. 2023;121:78–87. doi: 10.1016/j.neurobiolaging.2022.09.003.

32. Fröhlich S, Kutz DF, Müller K, Voelcker-Rehage C. Characteristics of Resting State EEG Power in 80+-Year-Olds of Different Cognitive Status. Frontiers in Aging Neuroscience. 2021;13. doi: 10.3389/fnagi.2021.675689.

33. Toepper M. Dissociating Normal Aging from Alzheimer’s Disease: A View from Cognitive Neuroscience. J Alzheimers Dis. 2017;57(2):331–52. doi: 10.3233/jad-161099.

34. Gao S, Hendrie HC, Hall KS, Hui S. The Relationships Between Age, Sex, and the Incidence of Dementia and Alzheimer Disease: A Meta-analysis. Archives of General Psychiatry. 1998;55(9):809–15. doi: 10.1001/archpsyc.55.9.809.

35. Qiu C, Fratiglioni L. Aging without dementia is achievable: current evidence from epidemiological research. Journal of Alzheimer’s Disease. 2018;62(3):933–42.

36. Nelson PT, Head E, Schmitt FA, Davis PR, Neltner JH, Jicha GA, et al. Alzheimer’s disease is not “brain aging”: neuropathological, genetic, and epidemiological human studies. Acta neuropathologica. 2011;121:571–87.

37. Miltiadous A, Tzimourta KD, Afrantou T, Ioannidis P, Grigoriadis N, Tsalikakis DG, et al. A Dataset of Scalp EEG Recordings of Alzheimer’s Disease, Frontotemporal Dementia and Healthy Subjects from Routine EEG. Data. 2023 doi: 10.3390/data8060095.

38. Delorme A, Sejnowski T, Makeig S. Enhanced detection of artifacts in EEG data using higher-order statistics and independent component analysis. Neuroimage. 2007;34(4):1443–9. doi: 10.1016/j.neuroimage.2006.11.004.

39. Miltiadous A, Tzimourta KD, Giannakeas N, Tsipouras MG, Afrantou T, Ioannidis P, Tzallas AT. Alzheimer’s Disease and Frontotemporal Dementia: A Robust Classification Method of EEG Signals and a Comparison of Validation Methods. Diagnostics (Basel). 2021;11(8). doi: 10.3390/diagnostics11081437.

40. Solomon Jr OM. PSD computations using Welch’s method. NASA STI/Recon Technical Report N. 1991;92:23584.

41. Kurlowicz L, Wallace M. The Mini-Mental State Examination (MMSE). J Gerontol Nurs. 1999;25(5):8–9. doi: 10.3928/0098-9134-19990501-08.

42. Beason-Held LL, Goh JO, An Y, Kraut MA, O’Brien RJ, Ferrucci L, Resnick SM. Changes in brain function occur years before the onset of cognitive impairment. J Neurosci. 2013;33(46):18008–14. doi: 10.1523/jneurosci.1402-13.2013.

43. Chouliaras L, O’Brien JT. The use of neuroimaging techniques in the early and differential diagnosis of dementia. Molecular Psychiatry. 2023;28(10):4084–97. doi: 10.1038/s41380-023-02215-8.

44. Ribarič S. Detecting Early Cognitive Decline in Alzheimer’s Disease with Brain Synaptic Structural and Functional Evaluation. Biomedicines. 2023 doi: 10.3390/biomedicines11020355.

45. Talwar P, Kushwaha S, Chaturvedi M, Mahajan V. Systematic review of different neuroimaging correlates in mild cognitive impairment and Alzheimer’s disease. Clinical neuroradiology. 2021;31(4):953–67.

46. Zhang H, Wang Y, Lyu D, Li Y, Li W, Wang Q, et al. Cerebral blood flow in mild cognitive impairment and Alzheimer’s disease: A systematic review and meta-analysis. Ageing research reviews. 2021;71:101450.

47. Sun J, Wang B, Niu Y, Tan Y, Fan C, Zhang N, et al. Complexity analysis of EEG, MEG, and fMRI in mild cognitive impairment and Alzheimer’s disease: a review. Entropy. 2020;22(2):239.

48. Jung J, Cloutman LL, Binney RJ, Lambon Ralph MA. The structural connectivity of higher order association cortices reflects human functional brain networks. Cortex. 2017;97:221–39. doi: 10.1016/j.cortex.2016.08.011.

49. Friedman NP, Robbins TW. The role of prefrontal cortex in cognitive control and executive function. Neuropsychopharmacology. 2022;47(1):72–89. doi: 10.1038/s41386-021-01132-0.

50. Takeuchi H, Taki Y, Sassa Y, Hashizume H, Sekiguchi A, Fukushima A, Kawashima R. Brain structures associated with executive functions during everyday events in a non-clinical sample. Brain Struct Funct. 2013;218(4):1017–32. doi: 10.1007/s00429-012-0444-z.

51. Ullman MT. Contributions of memory circuits to language: the declarative/procedural model. Cognition. 2004;92(1):231–70. doi: 10.1016/j.cognition.2003.10.008.

52. Busch RM, Yehia L, Blümcke I, Hu B, Prayson R, Hermann BP, et al. Molecular and subregion mechanisms of episodic memory phenotypes in temporal lobe epilepsy. Brain Commun. 2022;4(6):fcac285. doi: 10.1093/braincomms/fcac285.

53. Wiltgen BJ, Zhou M, Cai Y, Balaji J, Karlsson MG, Parivash SN, et al. The hippocampus plays a selective role in the retrieval of detailed contextual memories. Curr Biol. 2010;20(15):1336–44. doi: 10.1016/j.cub.2010.06.068.

54. Perry R, Hodges J. Differentiating frontal and temporal variant frontotemporal dementia from Alzheimer’s disease. Neurology. 2000;54(12):2277–84.

55. Kumfor F, Irish M, Hodges JR, Piguet O. Frontal and temporal lobe contributions to emotional enhancement of memory in behavioral-variant frontotemporal dementia and Alzheimer’s disease. Frontiers in behavioral neuroscience. 2014;8:225.

56. Berron D, van Westen D, Ossenkoppele R, Strandberg O, Hansson O. Medial temporal lobe connectivity and its associations with cognition in early Alzheimer’s disease. Brain. 2020;143(4):1233–48.

57. Prichep LS, John ER, Ferris SH, Reisberg B, Almas M, Alper K, Cancro R. Quantitative EEG correlates of cognitive deterioration in the elderly. Neurobiology of Aging. 1994;15(1):85–90. doi: 10.1016/0197-4580(94)90147-3.

58. Tedrus GM, Negreiros LM, Ballarim RS, Marques TA, Fonseca LC. Correlations Between Cognitive Aspects and Quantitative EEG in Adults With Epilepsy. Clinical EEG and Neuroscience. 2018;50(5):348–53. doi: 10.1177/1550059418793553.

59. Corsi-cabrera M, Herrera P, Malvido M. Correlation Between Eeg and Cognitive Abilities: Sex Differences. International Journal of Neuroscience. 1989;45(1-2):133–41. doi: 10.3109/00207458908986226.

60. Savva GM, Wharton SB, Ince PG, Forster G, Matthews FE, Brayne C. Age, neuropathology, and dementia. New England Journal of Medicine. 2009;360(22):2302–9.

61. Gonzales MM, Garbarino VR, Pollet E, Palavicini JP, Kellogg DL, Kraig E, Orr ME. Biological aging processes underlying cognitive decline and neurodegenerative disease. The Journal of clinical investigation. 2022;132(10).

62. Wu JW, Yaqub A, Ma Y, Koudstaal W, Hofman A, Ikram MA, et al. Biological age in healthy elderly predicts aging-related diseases including dementia. Scientific reports. 2021;11(1):15929.

63. Mecocci P, Boccardi V. The impact of aging in dementia: It is time to refocus attention on the main risk factor of dementia. Ageing research reviews. 2021;65:101210.

64. Fang X, Crumpler RF, Thomas KN, Mazique JN, Roman RJ, Fan F. Contribution of cerebral microvascular mechanisms to age-related cognitive impairment and dementia. Physiology international. 2022;109(1):20–30.

65. Ton AMM, Campagnaro BP, Alves GA, Aires R, Côco LZ, Arpini CM, et al. Oxidative stress and dementia in Alzheimer’s patients: effects of synbiotic supplementation. Oxidative medicine and cellular longevity. 2020;2020(1):2638703.

66. Ashleigh T, Swerdlow RH, Beal MF. The role of mitochondrial dysfunction in Alzheimer’s disease pathogenesis. Alzheimer’s & Dementia. 2023;19(1):333–42.

67. Sharma C, Kim S, Nam Y, Jung UJ, Kim SR. Mitochondrial dysfunction as a driver of cognitive impairment in Alzheimer’s disease. International Journal of Molecular Sciences. 2021;22(9):4850.

68. Karabiyik C, Frake RA, Park SJ, Pavel M, Rubinsztein DC. Autophagy in ageing and ageing-related neurodegenerative diseases. Ageing Neurodegener Dis. 2021;1(2).

69. Wang X-X, Zhang B, Xia R, Jia Q-Y. Inflammation, apoptosis and autophagy as critical players in vascular dementia. European Review for Medical & Pharmacological Sciences. 2020;24(18).

70. Lutshumba J, Nikolajczyk BS, Bachstetter AD. Dysregulation of systemic immunity in aging and dementia. Frontiers in Cellular Neuroscience. 2021;15:652111.

